# Neuropsychiatric Symptom Associations with *NOTCH3* Genotype in the United States CADASIL Consortium

**DOI:** 10.64898/2026.09.24.26363718

**Authors:** Megan R. Caruso, Barbara L. Fischer, William H. Adams, Deven K. Burks, Bianca Le, Arash Salardini, Jennifer J. Majersik, Stephen P. Salloway, Jamie L. Elliott, H. Jeremy Bockholt, Suman Jayadev, Michael D. Geschwind, José Biller, Helmi L. Lutsep, Sara J. Doyle, Jason D. Hinman, David S. Liebeskind, Imama A. Naqvi, Kelsey Eklund, Sudha Seshadri, Lisa C. Krishnamurthy, Myriam Fornage, Jane S. Paulsen, Edward D. Huey, the United States CADASIL Consortium

## Abstract

**Background and objectives:** Cerebral Autosomal Dominant Arteriopathy with Subcortical Infarcts and Leukoencephalopathy (CADASIL) is the most common monogenic cerebral small vessel disease. Though neuropsychiatric symptoms (NPS) of CADASIL profoundly impact function and quality of life, these symptoms remain understudied. This analysis characterizes the prevalence, severity, and profile of NPS among individuals with CADASIL-causing *NOTCH3* variants and non-carrier biological family controls in the United States CADASIL Consortium (USCC).

**Methods:** This cross-sectional analysis used multimodal baseline data from 12 enrollment sites across the country. Statistical analyses used nonparametric tests and regression models to compare clinical and neuropsychiatric outcomes across CADASIL risk tiers and controls, adjusting for age, sex, race, and education where appropriate. The primary outcome measure was the Neuropsychiatric Inventory Questionnaire (NPI-Q), an informant-reported measure which assesses 12 categories of NPS. NPI-Q item severity and distress were evaluated for individuals with confirmed cysteine-altering pathogenic *NOTCH3* variants and compared with controls.

**Results:** Four hundred fifty-nine USCC participants were included in the analytic cohort: 343 cases [256 high-risk (HR); 63 medium-risk (MR); 24 low-risk (LR)]; and 116 controls. Among participants with available NPI-Q data, at least one neuropsychiatric symptom was reported by 72.6% (n = 228/314) of cases compared with 53.2% (n = 58/109) of controls. Reported symptoms were more common among CADASIL cases than controls, occurring in 75.0%, 71.2%, and 72.7% of participants in the LR, MR, and HR groups, respectively, compared with 53.2% of controls. Compared with controls, participants in the MR group and HR group had higher NPI-Q severity scores. HR participants also had higher NPI-Q distress scores than controls.

**Discussion:** Our findings support the notion that NPS are an important clinical manifestation of CADASIL. Beyond previously reported symptoms, additional neuropsychiatric manifestations were identified. Given the higher burden and variability in the neuropsychiatric presentation of CADASIL, the relationships among neuropsychiatric, neurological, and patient-reported outcomes merit further investigation. The NPS profile may enhance the evaluation of the safety and efficacy of different treatments to target these symptoms for CADASIL patients.

Trial Registration Information: The study is registered at ClinicalTrials.gov (NCT05677880, 12/12/2022).

## Introduction

Cerebral Autosomal Dominant Arteriopathy with Subcortical Infarcts and Leukoencephalopathy (CADASIL) is the most common monogenic cause of cerebral small vessel disease (cSVD).^1–6^ In the general population, cSVD is a known cause of stroke, cognitive impairment, and disordered mood in older adults.^5^ Though rare, CADASIL can serve as a model for small vessel ischemic changes, with regard to numerous symptoms including neuropsychiatric symptoms.^1, 7^ While the disease course can be highly variable, recent work has described phenotypic variability across three tiers based on genotype, as originally proposed by Hack and colleagues in 2023.^8^ The onset of symptoms is typically observed in the third to fourth decade of life.^1, 7, 9^ Clinically, early symptoms may include behavioral changes and neuropsychiatric symptoms (NPS), which can exert a profound impact on function and quality of life.^9–11^ To date, however, much remains unknown about NPS in CADASIL.

In previous studies, psychiatric disturbances were reported to occur in up to one-third to one-half of persons with a CADASIL-causing *NOTCH3* variant, often manifesting initially as dysphoric mood,^9–13^ followed by apathy in later stages of the disease.^10, 11^ A Chinese cohort (n=78) identified irritability as the most frequently observed NPS, followed by depression and apathy.^10^ Other neuropsychiatric manifestations associated with CADASIL include manic episodes,^14–16^ obsessive-compulsive tendencies,^17^ and psychosis.^13, 18^ These findings are, however, drawn from case reports, retrospective studies, and small cohorts, limiting their generalizability to the broader disease population. Providers need a deeper understanding of NPS to help patients and families navigate the complex, and sometimes devastating, effects of this disease. By identifying representative symptoms and symptom profiles, clinicians may aid patients and families to set appropriate expectations, normalize their experiences, and manage challenging behaviors.

In this study, we utilize baseline data from the United States CADASIL Consortium (USCC) to describe the clinical and neuropsychiatric manifestations of CADASIL. We comprehensively examine the prevalence, severity, and profile of NPS across three gradations of genotype-derived risk. The primary goal of this analysis was to characterize the most common neuropsychiatric presentations of CADASIL by comparing Neuropsychiatric Inventory Questionnaire (NPI-Q) ratings across cases and non-carrier family members.

## Methods

### Study Design and Setting

This cross-sectional analysis was conducted using baseline data collected between October 2022 and January 2025 from the USCC. The USCC is a multicenter, longitudinal natural history study following participants across three visits, 18 months apart, at 12 enrollment sites in the United States. The study follows the cross-sectional extension of the Strengthening the Reporting of Observational Studies in Epidemiology (STROBE) reporting guidelines for observational research studies.^19^ The USCC’s design, setting, and detailed methods have been previously described.^20^

### Standard Protocol Approvals, Registrations, and Patient Consents

All versions of the USCC study protocol, recruitment materials, and site consent forms were approved by the Western Clinical Group (WCG). All participants and companions provided written informed consent before beginning any study-related procedures.

### Participant Selection and Cohort Definitions

Relevant eligibility criteria for participants included being an individual over the age of 18, a known or suspected personal or family history of CADASIL, and having a companion who knows them well (>3 hours a week of contact) and was willing to answer surveys about the participant. Exclusion criteria involved comorbid neurological conditions affecting the interpretability of findings, uncontrolled systemic diseases, and/or severe functional impairment defined as a modified Rankin scale (mRS) score >3 (i.e., indicating significant impairment of independence and mobility). For this analysis, we only included individuals with a pathogenic or likely pathogenic cysteine-altering *NOTCH3* variant and non-carrier family members. The National Centralized Repository for Alzheimer’s Disease and Related Dementias (NCRAD) processed and biobanked biospecimens, with central genetic testing conducted at the University of Wisconsin–Madison Next Generation Sequencing Division, as described elsewhere.^20^

### Clinical Assessments

Cognitive, behavioral, and functional impairments are quantified using the Clinical Dementia Rating (CDR) scale (Global Score and Sum of Boxes), modified Rankin Scale (mRS), 12-item World Health Organization Disability Assessment Schedule (WHODAS-12), Montreal Cognitive Assessment (MoCA), NPI-Q, and Starkstein Apathy Scale (SAS).^21–26^ USCC baseline findings for the CDR, mRS, WHODAS-12, and MoCA have been reported elsewhere.^20^

The focus of the present analysis is the NPI-Q, a widely used instrument that assesses 12 categories of NPS associated with Alzheimer’s disease (AD) and other neurodegenerative diseases.^26^ It is administered to the participant’s designated companion as part of a short informant interview that occurs within 30 days of the participant’s visit. Because many USCC participants have not yet experienced memory changes, a modification was made to the standardized administration guidelines. The phrase “based on changes that have occurred since the patient first began to experience memory problems” was omitted,^27, 28^ and informants were asked to respond instead based on their recent observations.

With the large scope of topics covered and its ease of use, the NPI-Q has merit in both clinical and research applications.^29^ One factor structure of NPI-Q, derived from responses collected at AD research centers, indicated that NPS generally cluster into four domains: mood/affective disturbance, behavioral activation, behavioral inhibition/somatic disturbance, and psychosis.^30, 31^

These domains provide context regarding the breadth of symptoms represented across USCC participants.

### Genetic Stratification and CADASIL Risk Tiers

*NOTCH3* variants were categorized into risk tiers: high-risk (HR), medium-risk (MR), and low-risk (LR), based on the specific epidermal growth factor-like repeat (EGFr) domain in which the variant is found. This stratification was modelled on Hack and coauthors’ genotype-phenotype risk classification system.^8^

### Statistical Methods

Baseline demographic and clinical characteristics were compared across control, LR, MR, and HR tiers. Categorical variables were compared using Fisher exact tests whereas continuous and ordinal variables were compared using Kruskal–Wallis tests. When omnibus Kruskal–Wallis tests were statistically significant, post hoc pairwise comparisons were performed using the Dwass–Steel–Critchlow–Fligner method, which controls the family-wise error rate across pairwise comparisons.^32–34^ For categorical variables with statistically significant Fisher exact tests, exact logistic regression was used for pairwise comparisons because of sparse cell counts. No additional adjustment for multiple testing across baseline characteristics was performed because the primary objective was descriptive characterization of the cohort.^35^

For the NPI-Q, informants’ responses were scored according to published guidelines, and the total severity and distress scores were calculated among participants with complete NPI-Q data. ^27^ Ordinal NPI-Q severity and caregiver distress ratings were initially compared across CADASIL risk tiers and controls using Kruskal–Wallis tests with Dwass–Steel–Critchlow-Fligner post hoc comparisons when appropriate. Associations between CADASIL risk tier and NPI-Q severity and distress summary scores were further evaluated using univariable and multivariable general linear models. Total NPI-Q symptom counts were evaluated using univariable and multivariable negative binomial regression models with a log link. Multivariable models adjusted *a priori* for age, sex, race, and years of education.

To evaluate the latent structure of NPS, a confirmatory factor analysis (CFA) was performed among cases using the prespecified four-factor model previously reported by González and coauthors.^30, 31^ The hypothesized model included latent factors representing psychosis (hallucinations, delusions), behavioral activation (agitation, irritability, disinhibition), internalizing symptoms (anxiety, depression), and somatic disturbance/behavioral suppression (apathy, appetite changes, abnormal nighttime behaviors). Consistent with the previously published model, elation and motor disturbance were not assigned to latent factors. Because NPI-Q responses are ordinal, model parameters were estimated using diagonally weighted least squares. Model fit was assessed using the root mean square error of approximation (RMSEA), standardized root mean square residual (SRMR), and comparative fit index (CFI). Standardized factor loadings were examined to evaluate the correspondence between observed NPI-Q domains and their hypothesized latent constructs. Missing data were limited and varied by measure; analyses used all available baseline data. All analyses were completed using SAS 9.4 (Cary, NC). All statistical tests were two-sided, and statistical significance was defined as *p* < .05.

### Data Availability

De-identified individual participant data that support the findings reported in this article will be available to qualified investigators upon request through the COINS data exchange service (https://coins.trendscenter.org).

## Results

**Figure 1** illustrates the cohort assembly. Of 560 enrolled participants, 101 (18.0%) were excluded because genome-wide association study data were unavailable at time of this analysis (9.8%) or because they carried a *NOTCH3* variant of uncertain significance (8.2%). The final analytic cohort included 459 participants (82.0%), comprising 343 CADASIL cases (74.7%) and 116 non-carrier family members (25.3%). Cases were further stratified into LR (24/343, 7.0%), MR (63/343, 18.4%), and HR (256/343, 74.6%) groups.

**Figure 1.**
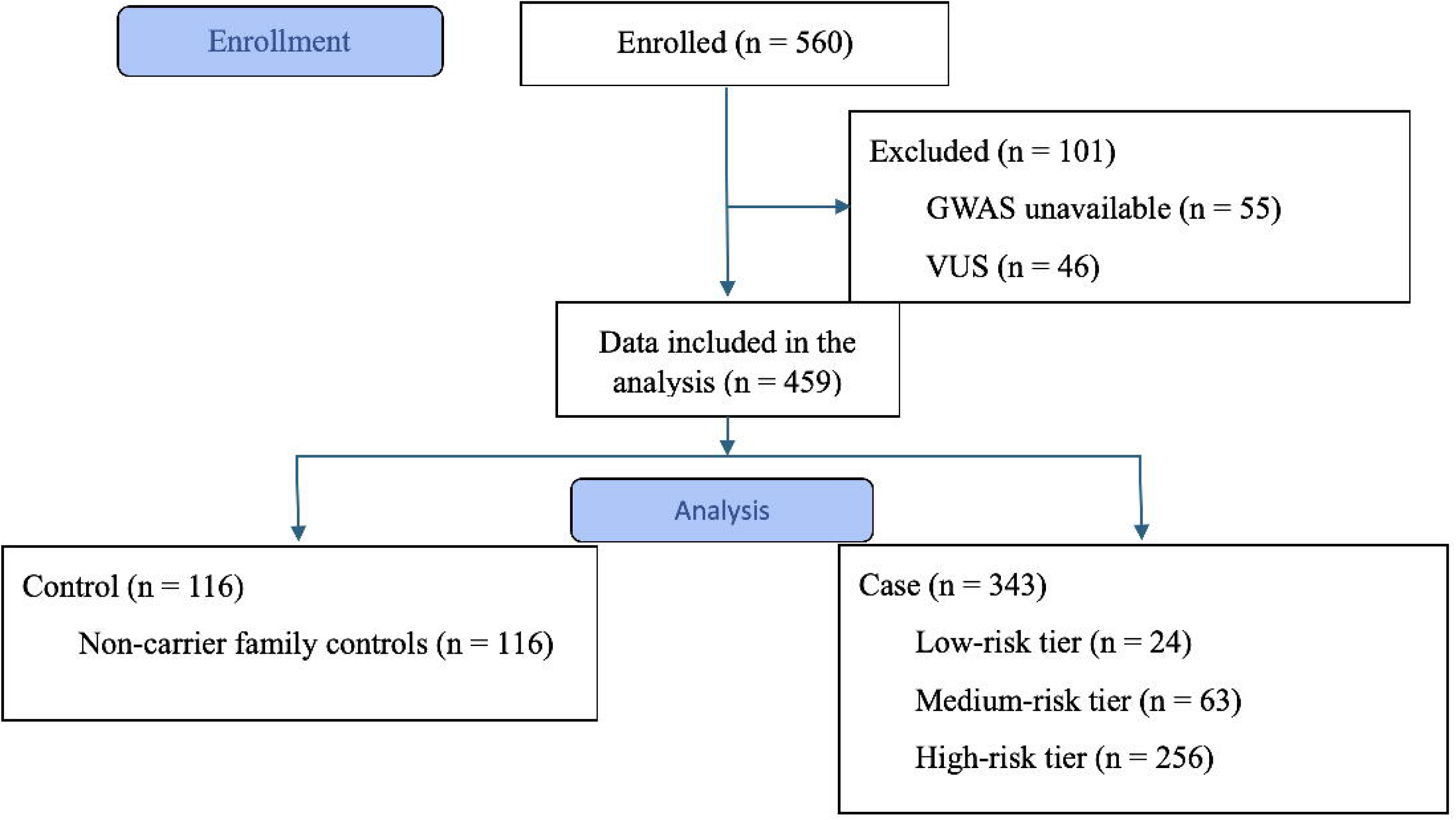
STROBE flow diagram.

Demographic and clinical characteristics are presented in **Table 1**. LR participants were more likely to identify as non-White when compared to controls (25.0% vs 6.1%, *p* = .02) and HR participants (25.0% vs 7.1%, *p* = .02). Additionally, MR participants were more likely to identify as Hispanic when compared to controls (19.0% vs 2.6%, *p* = .001) and HR Participants (19.0% vs 8.2%, *p* = .03). MR participants were also older than controls (*Mdn* = 55.4 vs 46.1; *p* = .03).

**Table 1.** Participant characteristics and clinical features by CADASIL risk tier.

|  | **CONTROL**<br>**(N=116)** | **LOW**<br>**(N=24)** | **MEDIUM**<br>**(N=63)** | **HIGH**<br>**(N=256)** | **Total**<br>**(N=459)** | ***p*** |
| --- | --- | --- | --- | --- | --- | --- |
| **Sex, n (%)** |  |  |  |  |  | .718<sup>1</sup> |
| Female | 76 (65.5%) | 16 (66.7%) | 41 (65.1%) | 154 (60.2%) | 287 (62.5%) |  |
| Male | 40 (34.5%) | 8 (33.3%) | 22 (34.9%) | 102 (39.8%) | 172 (37.5%) |  |
| **Race, n (%)** |  |  |  |  |  | .018<sup>1</sup> |
| non-White | 7 (6.1%) | 6 (25.0%) | 8 (12.9%) | 18 (7.1%) | 39 (8.6%) |  |
| White | 108 (93.9%) | 18 (75.0%) | 54 (87.1%) | 234 (92.9%) | 414 (91.4%) |  |
| Missing | 1 | 0 | 1 | 4 | 6 |  |
| **Ethnicity, n (%)** |  |  |  |  |  | .002<sup>1</sup> |
| Not Hispanic or Latino | 113 (97.4%) | 21 (87.5%) | 51 (81.0%) | 235 (91.8%) | 420 (91.5%) |  |
| Hispanic or Latino | 3 (2.6%) | 3 (12.5%) | 12 (19.0%) | 21 (8.2%) | 39 (8.5%) |  |
| **Age** |  |  |  |  |  | .030<sup>2</sup> |
| N (Missing) | 116 (0) | 24 (0) | 63 (0) | 256 (0) | 459 (0) |  |
| Median (IQR) | 46.1 (33.0,<br>57.8) | 46.0 (39.4,<br>54.7) | 55.4 (43.2,<br>65.2) | 48.8 (38.8,<br>58.9) | 47.9 (38.7,<br>59.1) |  |
| **Years of education** |  |  |  |  |  | .064<sup>2</sup> |
| N (Missing) | 116 (0) | 24 (0) | 63 (0) | 256 (0) | 459 (0) |  |
| Median (IQR) | 16.0 (15.0,<br>18.0) | 15.5 (14.0,<br>17.0) | 16.0 (13.0,<br>18.0) | 16.0 (14.0,<br>16.0) | 16.0 (14.0,<br>17.0) |  |
| **Prior Stroke, n (%)** |  |  |  |  |  | <.001<sup>1</sup> |
| No | 114 (98.3%) | 19 (79.2%) | 43 (69.4%) | 146 (57.9%) | 322 (70.9%) |  |
| Yes | 2 (1.7%) | 5 (20.8%) | 19 (30.6%) | 106 (42.1%) | 132 (29.1%) |  |
| Missing or unknown | 0 | 0 | 1 | 4 | 5 |  |
| **Prior Transient Ischemic**<br>**Attack (TIA), n (%)** |  |  |  |  |  | **<.001<sup>1</sup>** |
| No | 111 (95.7%) | 17 (70.8%) | 42 (66.7%) | 156 (62.2%) | 326 (71.8%) |  |
| Yes | 5 (4.3%) | 7 (29.2%) | 21 (33.3%) | 95 (37.8%) | 128 (28.2%) |  |
| Missing or unknown | 0 | 0 | 0 | 5 | 5 |  |
| **Age at first stroke or TIA** |  |  |  |  |  | **.527<sup>2</sup>** |
| N (Missing or not applicable) | 6 (110) | 9 (15) | 25 (38) | 149 (107) | 189 (270) |  |
| Median (IQR) | 48.0 (41.0,<br>53.0) | 43.0 (36.0,<br>45.0) | 48.0 (35.0,<br>53.0) | 43.0 (35.0,<br>50.0) | 43.0 (35.0,<br>51.0) |  |
| **History of migraine, n (%)** |  |  |  |  |  | **.002<sup>1</sup>** |
| No | 47 (40.5%) | 12 (50.0%) | 21 (33.3%) | 62 (24.3%) | 142 (31.0%) |  |
| Yes | 69 (59.5%) | 12 (50.0%) | 42 (66.7%) | 193 (75.7%) | 316 (69.0%) |  |
| Missing | 0 | 0 | 0 | 1 | 1 |  |
| **History of migraine with aura,**<br>**n (%)** |  |  |  |  |  | **.001<sup>1</sup>** |
| No | 34 (49.3%) | 5 (41.7%) | 15 (35.7%) | 46 (24.0%) | 100 (31.7%) |  |
| Yes | 35 (50.7%) | 7 (58.3%) | 27 (64.3%) | 146 (76.0%) | 215 (68.3%) |  |
| Missing or not applicable | 47 | 12 | 21 | 64 | 144 |  |
| **Migraine frequency, n (%)** |  |  |  |  |  | **.209<sup>2</sup>** |
| Never | 2 (2.9%) | 0 (0.0%) | 2 (4.8%) | 2 (1.0%) | 6 (1.9%) |  |
| Rare | 35 (50.7%) | 3 (25.0%) | 16 (38.1%) | 78 (40.6%) | 132 (41.9%) |  |
| Occasional | 7 (10.1%) | 2 (16.7%) | 3 (7.1%) | 39 (20.3%) | 51 (16.2%) |  |
| Frequent | 24 (34.8%) | 7 (58.3%) | 15 (35.7%) | 62 (32.3%) | 108 (34.3%) |  |
| Persistent | 1 (1.4%) | 0 (0.0%) | 6 (14.3%) | 11 (5.7%) | 18 (5.7%) |  |
| Missing or not applicable | 47 | 12 | 21 | 64 | 144 |  |
| **Confusional episode or**<br>**encephalopathy, n (%)** |  |  |  |  |  | .135<sup>1</sup> |
| No | 115 (99.1%) | 23 (95.8%) | 60 (95.2%) | 241 (94.5%) | 439 (95.9%) |  |
| Yes | 1 (0.9%) | 1 (4.2%) | 3 (4.8%) | 14 (5.5%) | 19 (4.1%) |  |
| Missing | 0 | 0 | 0 | 1 | 1 |  |
| **Mild Cognitive Impairment**<br>**(MCI), n (%)** |  |  |  |  |  | <.001<sup>1</sup> |
| No | 116 (100.0%) | 23 (95.8%) | 55 (87.3%) | 230 (90.2%) | 424 (92.6%) |  |
| Yes | 0 (0.0%) | 1 (4.2%) | 8 (12.7%) | 25 (9.8%) | 34 (7.4%) |  |
| Missing | 0 | 0 | 0 | 1 | 1 |  |
| **Dementia, n (%)** |  |  |  |  |  | .202<sup>1</sup> |
| No | 116 (100.0%) | 24 (100.0%) | 61 (96.8%) | 247 (96.9%) | 448 (97.8%) |  |
| Yes | 0 (0.0%) | 0 (0.0%) | 2 (3.2%) | 8 (3.1%) | 10 (2.2%) |  |
| Missing | 0 | 0 | 0 | 1 | 1 |  |
| **Depression, n (%)** |  |  |  |  |  | .321<sup>1</sup> |
| No | 72 (62.1%) | 14 (58.3%) | 30 (47.6%) | 144 (56.5%) | 260 (56.8%) |  |
| Yes | 44 (37.9%) | 10 (41.7%) | 33 (52.4%) | 111 (43.5%) | 198 (43.2%) |  |
| Missing | 0 | 0 | 0 | 1 | 1 |  |
| **Anxiety, n (%)** |  |  |  |  |  | .398<sup>1</sup> |
| No | 76 (65.5%) | 16 (66.7%) | 37 (58.7%) | 145 (56.9%) | 274 (59.8%) |  |
| Yes | 40 (34.5%) | 8 (33.3%) | 26 (41.3%) | 110 (43.1%) | 184 (40.2%) |  |
| Missing | 0 | 0 | 0 | 1 | 1 |  |
| **Obsessive-compulsive disorder, n (%)** |  |  |  |  |  | **.001<sup>1</sup>** |
| No | 116 (100.0%) | 24 (100.0%) | 57 (90.5%) | 252 (98.8%) | 449 (98.0%) |  |
| Yes | 0 (0.0%) | 0 (0.0%) | 6 (9.5%) | 3 (1.2%) | 9 (2.0%) |  |
| Missing | 0 | 0 | 0 | 1 | 1 |  |
| **Modified Rankin Scale score, n (%)** |  |  |  |  |  | **<.001<sup>2</sup>** |
| No symptoms | 86 (74.1%) | 12 (50.0%) | 23 (36.5%) | 82 (32.0%) | 203 (44.2%) |  |
| No significant disability | 25 (21.6%) | 7 (29.2%) | 19 (30.2%) | 97 (37.9%) | 148 (32.2%) |  |
| Slight disability | 2 (1.7%) | 3 (12.5%) | 16 (25.4%) | 40 (15.6%) | 61 (13.3%) |  |
| Moderate disability | 0 (0.0%) | 1 (4.2%) | 3 (4.8%) | 21 (8.2%) | 25 (5.4%) |  |
| mRS < 4; exact score unavailable<sup>a</sup> | 3 (2.6%) | 1 (4.2%) | 2 (3.2%) | 16 (6.3%) | 22 (4.8%) |  |

<sup>1</sup>Fisher Exact *p*-value; <sup>2</sup>Kruskal-Wallis *p*-value; <sup>a</sup>For the modified Rankin Scale (mRS), exact scores were unavailable for 22 participants; these participants were known only to have an mRS < 4 and are shown descriptively but were excluded from the hypothesis test for mRS.

Cerebrovascular disease was more common among participants with pathogenic or likely pathogenic *NOTCH3* variants. Compared with controls, participants in the LR, MR, and HR groups were more likely to report physician-diagnosed stroke (20.8%, 30.6%, and 42.1%, respectively, vs 1.7%; all *p* < .05) and transient ischemic attack (TIA) (29.2%, 33.3%, and 37.8%, respectively, vs 4.3%; all *p* < .05). Migraine was more common among HR participants when compared to controls (75.7% vs 59.5%, *p* = .003) and LR participants (75.7% vs 50.0%, *p* = .02). Of those participants with a history of migraine, HR participants were also more likely than controls to report experiencing migraine with aura (76.0% vs 50.7%, p < .001).

With respect to NPS history, MR and HR participants were more likely than controls to report mild cognitive impairment (12.7% and 9.8%, respectively, vs 0.0%; both *p* < .001). Obsessive-compulsive disorder was also more commonly diagnosed in the MR group (n = 6/63 or 9.5%) when compared to controls (n = 0/116 or 0.0%; *p* = .002) and those in the HR group (n = 3/255 or 1.2%; *p* = .005).

Functional disability increased across CADASIL risk tiers. Compared with controls, mRS scores were significantly worse among participants in the LR (*z* = 2.6, *p* = .04), MR (*z* = 5.7, *p* < .001), and HR groups (*z* = 7.7, *p* < .001).

Detailed distributions of NPI-Q severity and caregiver distress ratings are shown in **Figure 2A– C** and **Supplementary Table S1**. Among participants with available data, at least one NPS was reported in 67.6% (n = 286/423) of the overall cohort. NPS prevalence was not significantly different between LR participants and controls (75.0% vs 53.2%; *p* = .08). However, when compared with controls, NPS prevalence was significantly higher in the MR tier (71.2% vs 53.2%; *p* = .03) and HR tier (72.7% vs 53.2%; *p* = .001).

**Figure 2A-C.**
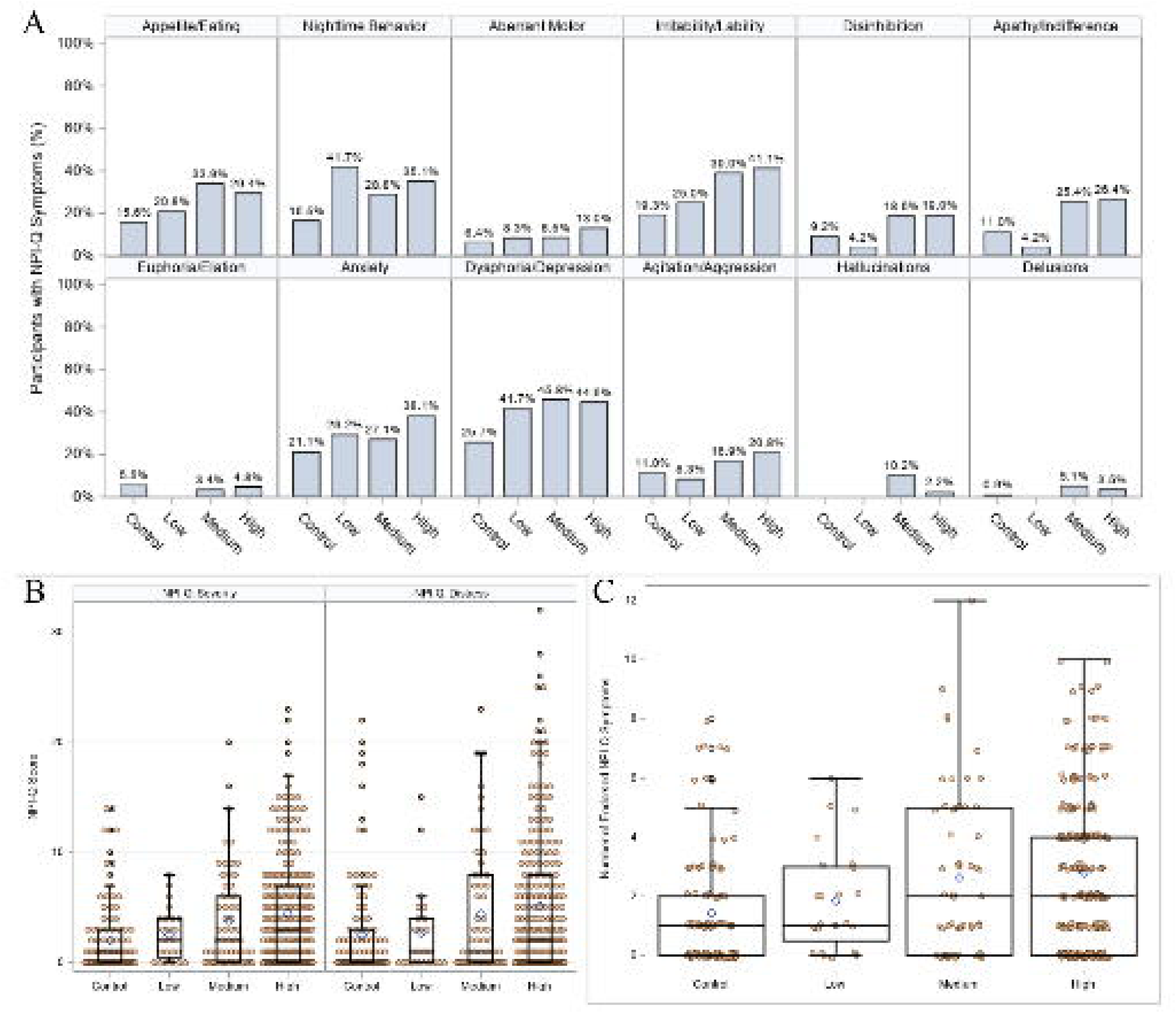
**A.** Distribution of NPI-Q responses by CADASIL risk tier. **B.** Distribution of NPI-Q scores by CADASIL risk tier. **C.** Number of endorsed NPI-Q symptoms by CADASIL risk tier.

NPS profiles also differed across risk tiers. HR participants were reported to exhibit both greater symptom severity and caregiver distress across multiple domains, including depression, anxiety, apathy, irritability, and appetite/eating disturbance (all p < .05). HR participants were also reported to have greater nighttime behavior disturbance severity (*p* = .002), but not caregiver distress scores. Relative to controls, MR participants were reported to demonstrate greater severity of depression, irritability, and appetite/eating disturbance (all *p* < .05). Hallucination severity and resulting caregiver distress were also observed to be greater in the MR group (both *p* < .05) although these differences were driven by 6 of 59 MR participants with available data, compared with none of the 109 controls with available data. LR participants were reported to have greater nighttime behavioral disturbance severity than controls (p = .047). Although the omnibus test for disinhibition severity was significant, no pairwise comparisons reached statistical significance.

Overall NPS burden increased with CADASIL risk tier. Compared with controls (*Mdn* = 1.0, IQR: 0.0 – 3.0), participants in the MR group (*Mdn* = 2.0, IQR: 0.0 – 6.0; *p* = .02) and HR group (*Mdn* = 3.0, IQR: 0.0 – 7.0; *p* < .001) obtained higher NPI-Q severity scores. HR participants also obtained higher NPI-Q caregiver distress scores than controls (*Mdn* = 2.0 vs 0.0; *p* < .001).

In multivariable regression models adjusting for age, sex, race, and education, HR participants obtained higher NPI-Q severity scores (β = 1.99, 95% CI 1.05 – 2.94; *p* < .001) and distress scores (β = 2.03, 95% CI 0.76 – 3.31; *p* = .002) relative to controls (**Table 2**).

**Table 2.** Associations Between CADASIL Risk Tier and NPI-Q scores.

|  | **Valid**<br>**N** | **Unadjusted $\beta$**<br>**(95% CI)** | ***p*** | **Adjusted $\beta$ (95%**<br>**CI)** | ***p*** |
| --- | --- | --- | --- | --- | --- |
| **NPI-Q Severity Score** |  |  |  |  |  |
| Group | 422 |  | <.001<sup>a</sup> |  | .001<sup>a</sup> |
| *High vs Control* |  | 2.42 (1.44, 3.40) | <.001 | 1.99 (1.05, 2.94) | <.001 |
| *Medium vs Control* |  | 1.78 (0.41, 3.14) | .011 | 1.02 (-0.30, 2.34) | .130 |
| *Low vs Control* |  | 0.52 (-1.38, 2.43) | .590 | 0.19 (-1.63, 2.02) | .838 |
| Male vs Female | 422 | 0.24 (-0.64, 1.11) | .597 | 0.38 (-0.44, 1.19) | .366 |
| Non-White vs White | 416 | 0.29 (-1.19, 1.76) | .703 | 0.69 (-0.71, 2.09) | .336 |
| Hispanic vs Non-Hispanic | 422 | 0.39 (-1.11, 1.88) | .612 |  |  |
| Education (per year) | 422 | -0.47 (-0.64, -0.29) | <.001 | -0.43 (-0.60, -0.26) | <.001 |
| Age (per year) | 422 | 0.05 (0.02, 0.08) | .001 | 0.05 (0.02, 0.08) | .001 |
| **NPI-Q Distress Score** |  |  |  |  |  |
| Group | 421 |  | .001<sup>a</sup> |  | .007<sup>a</sup> |
| *High vs Control* |  | 2.55 (1.24, 3.86) | <.001 | 2.03 (0.76, 3.31) | .002 |
| *Medium vs Control* |  | 1.87 (0.05, 3.69) | .044 | 0.92 (-0.85, 2.69) | .307 |
| *Low vs Control* |  | 0.13 (-2.41, 2.67) | .920 | -0.34 (-2.79, 2.10) | .783 |
| Male vs Female | 421 | 0.15 (-1.01, 1.30) | .803 | 0.35 (-0.74, 1.45) | .525 |
| Non-White vs White | 415 | 0.85 (-1.08, 2.78) | .387 | 1.35 (-0.53, 3.23) | .158 |
| Hispanic vs Non-Hispanic | 421 | 0.88 (-1.10, 2.85) | .383 |  |  |
| Education (per year) | 421 | -0.51 (-0.74, -0.28) | <.001 | -0.46 (-0.69, -0.23) | <.001 |
| Age (per year) | 421 | 0.06 (0.02, 0.10) | .003 | 0.06 (0.02, 0.09) | .005 |
| Note: CI = confidence interval; NPI-Q = Neuropsychiatric Inventory Questionnaire. Valid N = The number of participants used to compute the unadjusted estimates. The number of participants used to compute the adjusted estimates was 416 (for the severity sub-score) and 415 (for the distress sub-score). <sup>a</sup>Omnibus Type III *p*-value. |  |  |  |  |  |

Consistent with these findings, total NPI-Q symptom counts differed across CADASIL risk tiers and controls (*p* < .001). Median (IQR) symptom counts were 1.0 (0.0 – 2.0) among controls, 1.0 (0.5 – 3.0) in the LR group, 2.0 (0.0 – 5.0) in the MR group, and 2.0 (0.0 – 4.0) in the HR group. In pairwise comparisons, symptom counts were significantly higher in the MR (*z* = 3.0, *p* = .02) and HR (*z* = 4.9, *p* < .001) tiers than in controls whereas the LR tier did not differ significantly from controls (*z* = 1.7, *p* = .32). After adjusting for age, sex, race, and education, risk tier remained significantly associated with NPI-Q symptom count (*p* = .002). Compared with controls, participants in the HR tier had a 72% higher expected symptom count [rate ratio (*RR*) = 1.72, 95% CI: 1.31 – 2.25; *p* < .001)], and those in the MR tier had a 53% higher expected symptom count (*RR* = 1.53, 95% CI: 1.06 – 2.21; *p* = .02). The LR tier did not differ significantly from controls (*RR* = 1.31, 95% CI: 0.78 – 2.19; *p* = .30). Detailed results are presented in **Table 3**.

**Table 3.** Associations Between CADASIL Risk Tier and the count of reported NPI-Q items.

|  | **Valid**<br>**N** | **Unadjusted RR**<br>**(95% CI)** | ***p*** | **Adjusted RR**<br>**(95% CI)** | ***p*** |
| --- | --- | --- | --- | --- | --- |
| **Group** | 423 |  | <.001<sup>a</sup> |  | .002<sup>a</sup> |
| High vs Control |  | 1.95 (1.48, 2.57) | <.001 | 1.72 (1.31, 2.25) | <.001 |
| Medium vs Control |  | 1.85 (1.28, 2.68) | .001 | 1.53 (1.06, 2.21) | .022 |
| Low vs Control |  | 1.29 (0.76, 2.19) | .349 | 1.31 (0.78, 2.19) | .304 |
| **Male vs Female** | 423 | 1.06 (0.84, 1.34) | .600 | 1.09 (0.87, 1.35) | .448 |
| **Non-White vs White** | 417 | 1.02 (0.69, 1.52) | .913 | 1.03 (0.70, 1.50) | .895 |
| **Hispanic vs Non-Hispanic** | 423 | 1.13 (0.76, 1.67) | .555 |  |  |
| **Education** (per year) | 423 | 0.88 (0.84, 0.93) | <.001 | 0.90 (0.85, 0.94) | <.001 |
| **Age** (per year) | 423 | 1.01 (1.004, 1.02) | .005 | 1.01 (1.002, 1.02) | .012 |
| Note: RR = Ratio Ratio; CI = confidence interval. Valid N = The number of participants used to compute the unadjusted estimates. The number of participants used to compute the adjusted estimates was 417. <sup>a</sup>Omnibus Type III p-value. |  |  |  |  |  |

The previously reported four-factor NPI-Q model also demonstrated acceptable overall fit among cases (**Table 4**, **Supplemental Figure S1**). Model fit indices supported the hypothesized structure [χ² (df = 29) = 58.0 (*p* = .001); CFI = 0.96; SRMR = 0.04; RMSEA = 0.057, 90% CI 0.035 – 0.078)]. Standardized factor loadings were statistically significant for all NPI-Q domains (all *p* < .05) and ranged from 0.39 to 0.73. The strongest loading was observed for apathy/indifference (0.73) whereas the weakest loading was observed for delusions (0.39).

**Table 4.** Confirmatory factor analysis of the NPI-Q.

|  | **Standardized Loading** | **Reliability** | **Variance Extracted** |
| --- | --- | --- | --- |
| **Psychosis** |  | 0.40<sup>a</sup> | 0.26 |
| Delusions | 0.39 | 0.15 |  |
| Hallucinations | 0.60 | 0.36 |  |
| **Behavioral Activation** |  | 0.71<sup>a</sup> | 0.45 |
| Agitation/Aggression | 0.59 | 0.34 |  |
| Disinhibition | 0.71 | 0.50 |  |
| Irritability/Lability | 0.71 | 0.50 |  |
| **Internalizing Symptoms** |  | 0.53<sup>a</sup> | 0.36 |
| Depression/Dysphoria | 0.64 | 0.41 |  |
| Anxiety | 0.56 | 0.31 |  |
| **Somatic Disturbance/Behavioral Suppression** |  | 0.67<sup>a</sup> | 0.40 |
| Apathy/Indifference | 0.73 | 0.53 |  |
| Sleep and Night-time disorders | 0.63 | 0.39 |  |
| Appetite and eating disorders | 0.54 | 0.29 |  |
| Note: Estimates are among cases with complete NPI-Q responses (N = 314). <sup>a</sup>Denotes composite reliability. Fit statistics: $\chi^2_{df=29} = 58.0$ ($p = .001$); Bentler Comparative Fit Index (CFI) = 0.958; Standardized Root Mean Square Residual (SRMR) = 0.042. Root Mean Square Error of Approximation (RMSEA) = 0.057 (90% CI: 0.035 to 0.078). | | | |

## Discussion

Our findings suggest that NPS represent an important and prominent clinical feature of CADASIL. These results expand upon the results of Bai and coauthors,^10^ extending them to a large North American cohort, case-controlled by non-carrier family members and stratified by *NOTCH3* genotype risk. To our knowledge, our study comprises the largest, focused investigation of NPS in CADASIL stratifying risk according to Hack and coauthors.^8^ In addition to the historically recognized symptoms of irritability, apathy, and depression, USCC participants were observed to demonstrate changes in sleep behaviors, disinhibition, anxiety, and appetite/eating behaviors. As expected, compared to controls, HR participants were reported to exhibit significantly greater frequency and severity of NPS; those in LR and MR groups were noted to display lower severity of NPS than the HR group, which were at times indistinguishable from controls. Notably, the relationship between NPS and other symptoms of CADASIL merits further investigation given the variability of NPS presentation across risk tiers despite consistently reported neurological symptoms and stroke in all three tiers. While unspecific to CADASIL, the presence of NPS may assist with diagnostic clarification, raising suspicion for the disorder in patients who have not received a clinical diagnosis. Taken together, NPS are frequently reported among CADASIL patients, and they warrant further investigation as well as timely intervention.

In this cohort spanning presymptomatic to moderate disease, NPS were reported at a high prevalence across the cohort: 72.6% of persons with a CADASIL-causing *NOTCH3* variant and 53.2% of non-carrier family controls were reported to exhibit at least one NPS. Of interest, NPS, such as mood changes, have been shown to occur more commonly in families with autosomal dominant neurodegenerative diseases than in the general public, regardless of carrier status.^36, 37^ Notably, across the cohort, a higher percentage of cases were reported to display NPS than cerebrovascular events such as stroke and TIA, which are hallmark features of CADASIL. Furthermore, across all cases, there was a similar rate of any reported NPS to migraine, another characteristic symptom of the disease; associations between migraine and NPS have been previously reported.^38^

Analysis of the types of symptoms described in CADASIL cases versus controls revealed both expected and unexpected results. Consistent with the existing literature, in the present study, depression/dysphoria was the most frequently reported symptom among carriers (n = 140/314 or 44.6%), followed by irritability/lability (n = 124/314 or 39.5%) and anxiety (n = 111/314 or 35.4%), While apathy was less prevalent (n = 77/314 or 24.5%) in our cohort than reported elsewhere,^11^ it was still observed in cases more frequently than in controls. Potential explanations for this finding could be due to our cohort’s relatively younger age and lower degree of functional impairment, with both potentially serving as proxies for lower disease burden. Unexpected and previously under-reported symptoms in our cohort’s cases included non-affective symptoms such as aberrant sleep/nighttime behaviors (n = 108/314 or 34.4%), appetite/eating behavioral changes (n = 93/314 or 29.6%), disinhibition (n = 56/314 or 17.8%), and hallucinations (n = 11/314 or 3.5%).

While NPS in CADASIL have historically been described as falling within the mood/affective disturbance domain of the NPI-Q, the participants in our cohort were observed to exhibit symptoms across all four domains. As such, our results suggest that the NPS presentation of CADASIL is extensive, reframing our understanding to encompass both a high prevalence and a wide range of NPS types. These findings are critically relevant both to the patients and families who struggle to navigate the neurobehavioral changes and to the clinicians diagnosing CADASIL patients and managing their NPS.

Broadly, NPS burden increased with higher CADASIL risk tier. HR participants obtained significantly higher total NPI-Q severity and informant distress scores than controls. While MR participants were reported to have higher unadjusted NPI-Q severity scores than controls, this association attenuated after adjustment for covariates. Despite exhibiting significantly higher rates of other CADASIL symptoms, like stroke and migraine, LR participants did not obtain an increased total NPI-Q severity score compared to controls. However, the risk tiers may not denote a clear stepwise increase in disease burden, raising the possibility that different EGFr-based tiers may be associated with distinct clinical profiles.

While HR participants were reported to have the highest neuropsychiatric disease burden across all groups, obtaining the greatest total severity scores, they also were reported to have significantly higher scores than controls in half of the individual items. Concomitantly, participants in the HR group exhibited the greatest severity of cerebrovascular disease, with the highest prevalence of migraine, prior stroke, and TIA. This group also demonstrated the poorest performance on the CDR and the poorest functional performance on the mRS.^20^ These findings raise the possibility that higher genetic risk may be associated with higher levels of vascular disease burden and NPS, but these were not specifically investigated here. Future analyses should evaluate these relationships while controlling confounds such as sleep duration, stress, lifestyle factors, etc.

Several unexpected findings were also observed in the LR and MR groups. For example, LR participants were observed to have the highest severity of nighttime behaviors across all groups. MR participants were the only case group to exhibit a significantly higher severity of hallucinations than controls. However, given the limited sample sizes in the LR (n = 24) and MR (n = 63) tiers, these findings should be interpreted with caution.

While several of the symptoms observed in the USCC were consistent with typical CADASIL^5, 7, 9, 10^ and vascular cognitive impairment presentation,^39, 40^ the breadth and depth of NPS reported in this cohort were unexpected, particularly given lower rates of other disease features, like cerebrovascular events and cognitive impairment. Indeed, although there was a significantly higher prevalence of self-reported mild cognitive impairment (MCI) diagnoses in cases than controls, relatively few participants met criteria for either MCI or dementia.^20^ Yet the breadth and clustering of symptoms in our cohort were similar to those reported among individuals with AD and mixed dementias.^41^ The CFA for Gonzales’ National Alzheimer’s Coordinating Center (NACC) dataset-derived NPI-Q four-factor cluster model demonstrated good fit for this cohort.^31^ This is consistent with the notion that mixed etiology, including vascular contribution, is common among individuals with cognitive decline.^42^ Recent comparison of NPS profiles in vascular disease with AD and other neurodegenerative syndromes revealed some overlap in symptoms particularly early in the disease course.^43^

There may be several possible explanations for the unexpectedly high prevalence and broad range of observed NPS in individuals with CADASIL-causing *NOTCH3* variants, of which some may be inherent to neurologic disease. First, CADASIL cases may experience a high level of NPS in early disease stages as a consequence of lifelong, progressive vascular disease burden.^7^ Alternatively, the upstream influences of specific genetic variants associated with CADASIL may independently influence and exacerbate NPS. These hypotheses could be evaluated in future studies by longitudinally investigating timing and associations of vascular events with NPS via neuroimaging. Additionally, it is possible that NPS occur more broadly among individuals in initial neurologic disease stages than has previously been reported. This hypothesis could be evaluated by carefully examining and comparing NPS prevalence and severity of participants with early-stage CADASIL to those in early stages of other life-altering neurologic diseases relative to healthy controls.

Another possible explanation implicates reaction to disease: the diagnosis of a familial life-altering disease such as CADASIL may be sufficient to elicit elevated levels of emotional symptom severity and distress regardless of etiology.^44^ Notably, living with and caring for a family member with chronic disease may also evoke distress, the magnitude of which may be investigated by comparing NPS in family members with and without autosomal dominant neurologic disease. Finally, comparing NPS in participants who were aware of their gene status with those who were not could help differentiate the relative contribution of genotype awareness to NPS manifestation. More thorough inquiry by clinicians and researchers is needed to better understand patient and family experience. As is common for NPS in other neurological disorders, the relationship between functional status and NPS in CADASIL is likely complex.^43^ Impaired functioning is a risk factor for certain psychiatric symptoms and disorders, such as depression, anxiety, and apathy. These data are, however, cross-sectional, meaning that the temporal relationship between NPS and functional impairment cannot be determined at this time. Further investigation into the relatedness of neurological and neuropsychiatric features of CADASIL should be pursued. It has been reported that neurological features cluster into three general phenotypes: those predominantly experiencing migraines attacks, those experiencing lacunar stroke and motor disability, and those experiencing cognitive impairment.^45^ Inclusion of NPS in other symptom models will provide a more robust understanding of how the full spectrum of CADASIL symptoms interrelate and co-occur. For example, integrated neuroimaging and neuropsychiatric data would anchor these NPS findings to more specific biomarkers.^10^

Uniquely, persons with pathogenic *NOTCH3* variants may also experience chronic pain, in the form of migraine pain and post-stroke pain, which can influence the emergence, severity, and presentation of NPS.^10, 46^ As NPS exert such a profound impact on functional abilities in both the aging and general population, investigation into the relationship between individual function, disability, and presenting NPS could elucidate the sequence of impairment, possibly providing opportunities for intervention. Many of these informant-reported NPS are thought to impact quality of life (QoL), and analyzing NPI-Q findings with participant-reported QoL outcomes are a necessary next step to further understand the specific nature of how these NPS affect functionality and QoL, as well as any discordance between informant-reported observations and participants’ lived experiences.

In our cohort, sleep disturbances emerged as novel and prominent NPS across CADASIL cases. Additional exploration into sleep disturbances in CADASIL is vital to help preserve wellness and a high QoL. For example, both CADASIL animal models and individuals with sleep disturbance have displayed impaired glymphatic function.^47, 48^ A multimodal exploration of sleep in CADASIL could determine whether sleep abnormalities represent a result of the disease process, a driving force in progression of disease, or a synergistic interaction of the two. Alternatively, sleep disorder and NPS may represent independent sequelae of stroke events rather than of the CADASIL disease process per se.

Our findings must be interpreted in the light of several limitations. Though we are reporting cross-sectional baseline data, our study design is longitudinal, and we intend to report follow-up conclusions as data become available. Additionally, kinship structure was unavailable for this baseline analysis because the genetic analyses needed to characterize familial relationships are ongoing. Consequently, analyses assumed independent observations. Future longitudinal analyses will account for familial relatedness once this information becomes available.

Furthermore, the USCC cohort is relatively young and unimpaired at baseline, meaning that we have not captured the full spectrum of disease severity. As a result, cases included asymptomatic individuals and other individuals who were unaware of their gene status, such that our results cannot be directly compared to cohorts entirely composed of established patients. Additionally, the quality of companion responses may be inconsistent as some companions were more effectively able to answer some questions than other companions. For example, a cohabitating significant other could more accurately report on severity of sleep disturbances than a friend living elsewhere.

Lastly, the NPI-Q is an imperfect measure of NPS. As noted above, we modified the NPI-Q instructions to improve its relevance to our cohort such that it was not administered in the standard format. Moreover, the NPI-Q lacks specificity on certain items because it was designed for brief administration that can be completed by informants themselves.^27^ Although significant differences emerged in appetite and sleep behaviors, neither question is specific or directional. From the NPI-Q alone, we cannot determine whether persons with pathogenic *NOTCH3* variants are sleeping more or less or at inappropriate times; similarly, we cannot identify the nature of appetite and eating changes.

To conclude, our findings indicate that individuals with a CADASIL-causing *NOTCH3* variant exhibit a high level of NPS burden across a broad domain of symptoms. If replicated, our results would reframe NPS as a hallmark or characteristic feature of CADASIL with direct clinical relevance, which contributes to the real-world struggles and likely reduced quality of life experienced by persons with CADASIL-causing *NOTCH3* variants and their families.

Specifically, with this new knowledge, clinicians can more effectively assist patients to manage disease symptoms and empower them to seek care or treatment. Additionally, with greater awareness of the patterns of NPS seen in CADASIL, researchers can better evaluate the safety and efficacy of different treatments that target these symptoms for CADASIL cases, and NPS could be considered as a potential clinical trial outcome target in studies of CADASIL. It will be critical for providers and clinicians to proactively assist patients and families to include management of NPS type, severity, and distress as part of their treatment plan. Consideration of NPS will be important to aid in earlier diagnosis and detection of CADASIL in those not presenting with other classical symptoms of CADASIL.

## Acknowledgment

This work was supported by the National Institutes of Health and Food and Drug Administration under award numbers RF1AG074608, R01AG082208, R01AG085602, and U01FD008399. This manuscript is the result of funding in whole or in part by the National Institutes of Health (NIH). It is subject to the NIH Public Access Policy. Through acceptance of this federal funding, NIH has been given a right to make this manuscript publicly available in PubMed Central upon the Official Date of Publication, as defined by NIH. The authors acknowledge the research staff of the United States CADASIL Consortium: https://cadasil.wisc.edu/wp-content/uploads/sites/2133/2026/08/CADASIL_Consortium_Acknowledgments.pdf. The authors thank the valuable contributions of study participants and families to advance CADASIL research.

## Supplemental Materials

**Supplemental Table S1.** *Neuropsychiatric Inventory Questionnaire (NPI-Q) Severity and Caregiver Distress Scores by CADASIL Risk Tier*

**Supplemental Figure S1**. *Confirmatory factor structure for the NPI-Q*. PSY = Psychosis. BEH = Behavioral Activation. INT = Internalizing Symptoms. SOM = Somatic Disturbance / Behavioral Suppression. HAL = Hallucinations. DEL = Delusions. AGI = Agitation/Aggression. IRR = Irritability/Lability. DIS = Disinhibition. ANX = Anxiety. DEP = Depression/Dysphoria. APA = Apathy/Indifference. APP = Appetite and eating disorders. NIT = Sleep and Night-time disorders.

**Supplemental File 1.** *Cross-sectional STROBE checklist*.

## Notes

### Competing Interest Statement

The authors have declared no competing interest.

### Author Declarations

Ethics committee/IRB of Western Clinical Group gave ethical approval for this work.

## References

1. Chabriat H, Joutel A, Dichgans M, Tournier-Lasserve E, Bousser MG. Cadasil. Lancet Neurol 2009;8:643–653.

2. Cho BPH, Auckland K, Graf S, Markus HS. Rare Sequence Variation Underlying Suspected Familial Cerebral Small-Vessel Disease. J Am Heart Assoc 2024;13:e035771.

3. Cho BPH, Nannoni S, Harshfield EL, et al. NOTCH3 variants are more common than expected in the general population and associated with stroke and vascular dementia: an analysis of 200 000 participants. J Neurol Neurosurg Psychiatry 2021;92:694–701.

4. Choi JC. Genetics of cerebral small vessel disease. J Stroke 2015;17:7–16.

5. Di Donato I, Bianchi S, De Stefano N, et al. Cerebral Autosomal Dominant Arteriopathy with Subcortical Infarcts and Leukoencephalopathy (CADASIL) as a model of small vessel disease: update on clinical, diagnostic, and management aspects. BMC Med 2017;15:41.

6. Ferguson AC, Thrippleton S, Henshall D, et al. Frequency and Phenotype Associations of Rare Variants in 5 Monogenic Cerebral Small Vessel Disease Genes in 200,000 UK Biobank Participants. Neurol Genet 2022;8:e200015.

7. Chabriat H, Bousser MG. Neuropsychiatric manifestations in CADASIL. Dialogues Clin Neurosci 2007;9:199–208.

8. Hack RJ, Gravesteijn G, Cerfontaine MN, et al. Three-tiered EGFr domain risk stratification for individualized NOTCH3-small vessel disease prediction. Brain 2023;146:2913–2927.

9. Chabriat H, Lesnik Oberstein S. Cognition, mood and behavior in CADASIL. Cereb Circ Cogn Behav 2022;3:100043.

10. Bai L, Yan H, Guo Y, et al. The prevalence of neuropsychiatric symptoms and correlation with MRI findings in CADASIL patients. Ann Clin Transl Neurol 2024;11:3010–3018.

11. Reyes S, Viswanathan A, Godin O, et al. Apathy: a major symptom in CADASIL. Neurology 2009;72:905–910.

12. Wang YM, Yu HY, Zhang L, Hu C. Comorbidity Between Recurrent Major Depressive Disorder and CADASIL: A Case Report With a Brief Review of Published Cases. Clin Case Rep 2025;13:e71350.

13. Valenti R, Poggesi A, Pescini F, Inzitari D, Pantoni L. Psychiatric disturbances in CADASIL: a brief review. Acta Neurol Scand 2008;118:291–295.

14. Kumar SK, Mahr G. CADASIL presenting as bipolar disorder. Psychosomatics 1997;38:397–398.

15. Wang J, Li J, Kong F, Lv H, Guo Z. Bipolar II disorder as the initial presentation of CADASIL: an underdiagnosed manifestation. Neuropsychiatr Dis Treat 2017;13:2175–2179.

16. Valenti R, Pescini F, Antonini S, et al. Major depression and bipolar disorders in CADASIL: a study using the DSM-IV semi-structured interview. Acta Neurol Scand 2011;124:390–395.

17. Canli D, Keskin M. CADASIL Syndrome Presenting as Obsessive-Compulsive Disorder: A Case Report. Noro Psikiyatr Ars 2025;62:90–93.

18. Harris JG, Filley CM. CADASIL: neuropsychological findings in three generations of an affected family. J Int Neuropsychol Soc 2001;7:768–774.

19. von Elm E, Altman DG, Egger M, et al. The Strengthening the Reporting of Observational Studies in Epidemiology (STROBE) statement: guidelines for reporting observational studies. J Clin Epidemiol 2008;61:344–349.

20. Paulsen JS, Bockholt HJ, Adams WH, et al. The United States CADASIL Consortium: Baseline Findings from a Natural History Study [preprint]. In: medRxiv, ed.2026.

21. Bonita R, Beaglehole R. Modification of Rankin Scale: Recovery of motor function after stroke. Stroke 1988;19:1497–1500.

22. Morris JC. The Clinical Dementia Rating (CDR): current version and scoring rules. Neurology 1993;43:2412–2414.

23. Nasreddine ZS, Phillips NA, Bedirian V, et al. The Montreal Cognitive Assessment, MoCA: a brief screening tool for mild cognitive impairment. J Am Geriatr Soc 2005;53:695–699.

24. Starkstein SE, Mayberg HS, Preziosi TJ, Andrezejewski P, Leiguarda R, Robinson RG. Reliability, validity, and clinical correlates of apathy in Parkinson’s disease. J Neuropsychiatry Clin Neurosci 1992;4:134–139.

25. World Health Organization. Global Physical Activity Questionnaire (GPAQ) Analysis Guide. Geneva: World Health Organization, 2010.

26. Cummings J. The Neuropsychiatric Inventory: Development and Applications. J Geriatr Psychiatry Neurol 2020;33:73–84.

27. Kaufer DI, Cummings JL, Ketchel P, et al. Validation of the NPI-Q, a brief clinical form of the Neuropsychiatric Inventory. J Neuropsychiatry Clin Neurosci 2000;12:233–239.

28. Cummings J. The Neuropsychiatric Inventory–Questionnaire: Background and Administration [online]. Available at: https://www.alz.org/getmedia/fac45de4-c45d-4227-8e32-1153b4cdf6f3/npiq-questionnaire.pdf. Accessed July 20.

29. Trzepacz PT, Saykin A, Yu P, et al. Subscale validation of the neuropsychiatric inventory questionnaire: comparison of Alzheimer’s disease neuroimaging initiative and national Alzheimer’s coordinating center cohorts. Am J Geriatr Psychiatry 2013;21:607–622.

30. Gonzalez DA, Finley JA, Patel SES, Soble JR. Practical Assessment of Neuropsychiatric Symptoms: Updated Reliability, Validity, and Cutoffs for the Neuropsychiatric Inventory Questionnaire. Am J Geriatr Psychiatry 2025;33:524–534.

31. Gonzalez DA, Resch ZJ, Obolsky MA, Soble JR. Comparing behavioral and psychological symptom structures on the Neuropsychiatric Inventory Questionnaire. Psychol Assess 2023;35:522–532.

32. Dwass M. Some k-Sample Rank-Order Tests. In: Olkin I, Ghurye SG, Hoeffding W, Madow WG, Mann HB, eds. Contributions to Probability and Statistics: Essays in Honor of Harold Hotelling. Stanford, CA: Stanford University Press, 1960: 198–202.

33. Steel RG. A rank sum test for comparing all pairs of treatments. Technometrics 1960;2:197–207.

34. Critchlow DE, Fligner MA. On distribution-free multiple comparisons in the one-way analysis of variance. Communications in Statistics – Theory and Methods 1991;20:127–139.

35. Hoffmann S, Lemster S, Collins G, et al. When to Adjust for Multiple Testing: A Unifying Guiding Principle. Biom J 2026;68:e70148.

36. O’Connor A, Rice H, Barnes J, et al. First presentation with neuropsychiatric symptoms in autosomal dominant Alzheimer’s disease: the Dominantly Inherited Alzheimer’s Network Study. J Neurol Neurosurg Psychiatry 2023;94:403–405.

37. Ramos C, Rios-Romenets S, Giraldo-Chica M, et al. Depression and anxiety disorder along life in familial Alzheimer’s disease in the API ADAD Colombia trial. Alzheimer’s & Dementia 2020;16:e041314.

38. Minen MT, Begasse De Dhaem O, Kroon Van Diest A, et al. Migraine and its psychiatric comorbidities. J Neurol Neurosurg Psychiatry 2016;87:741–749.

39. Santos AP, Lobo AOM, Santos C, et al. Behavioral and psychological symptoms of dementia associated with vascular impairment: a systematic review and meta-analysis. Braz J Psychiatry 2025;47:e20254225.

40. Tiel C, Sudo FK, Alves GS, et al. Neuropsychiatric symptoms in Vascular Cognitive Impairment: a systematic review. Dement Neuropsychol 2015;9:230–236.

41. Johnson DK, Watts AS, Chapin BA, Anderson R, Burns JM. Neuropsychiatric profiles in dementia. Alzheimer Dis Assoc Disord 2011;25:326–332.

42. Alafuzoff I, Libard S. Ageing-Related Neurodegeneration and Cognitive Decline. Int J Mol Sci 2024;25.

43. Bell Z, O’Connor MK, Moo LR. Neuropsychiatric presentations of common dementia syndromes: A concise review for primary care team members. J Am Geriatr Soc 2025;73:349–357.

44. van der Meer L, van Duijn E, Wolterbeek R, Tibben A. Offspring of a parent with genetic disease: childhood experiences and adult psychological characteristics. Health Psychol 2014;33:1445–1453.

45. Dupe C, Guey S, Biard L, et al. Phenotypic variability in 446 CADASIL patients: Impact of NOTCH3 gene mutation location in addition to the effects of age, sex and vascular risk factors. J Cereb Blood Flow Metab 2023;43:153–166.

46. Atee M, Morris T, Macfarlane S, Cunningham C. Pain in Dementia: Prevalence and Association With Neuropsychiatric Behaviors. J Pain Symptom Manage 2021;61:1215–1226.

47. Chong PLH, Garic D, Shen MD, Lundgaard I, Schwichtenberg AJ. Sleep, cerebrospinal fluid, and the glymphatic system: A systematic review. Sleep Med Rev 2022;61:101572.

48. Romay MC, Knutsen RH, Ma F, et al. Age-related loss of Notch3 underlies brain vascular contractility deficiencies, glymphatic dysfunction, and neurodegeneration in mice. J Clin Invest 2024;134.

