## Supplemental Table 1 for "Neuropsychiatric Symptom Associations with *NOTCH3* Genotype in the United States CADASIL Consortium"

**Supplemental Table S1.** *Neuropsychiatric Inventory Questionnaire (NPI-Q) Severity and Caregiver Distress Scores by CADASIL Risk Tier*

|  | **CONTROL (N=116)** | **LOW (N=24)** | **MEDIUM (N=63)** | **HIGH (N=256)** | **Total (N=459)** | ***p*** |
| --- | --- | --- | --- | --- | --- | --- |
| **Delusions Severity**, n (%) |  |  |  |  |  | .314^1^ |
| 0 | 108 (99.1%) | 24 (100.0%) | 56 (94.9%) | 223 (96.5%) | 411 (97.2%) |  |
| 1 | 1 (0.9%) | 0 (0.0%) | 3 (5.1%) | 6 (2.6%) | 10 (2.4%) |  |
| 2 | 0 (0.0%) | 0 (0.0%) | 0 (0.0%) | 2 (0.9%) | 2 (0.5%) |  |
| Missing | 7 | 0 | 4 | 25 | 36 |  |
| **Delusions Distress**, n (%) |  |  |  |  |  | .312^1^ |
| 0 | 108 (99.1%) | 24 (100.0%) | 56 (94.9%) | 223 (96.5%) | 411 (97.2%) |  |
| 1 | 0 (0.0%) | 0 (0.0%) | 1 (1.7%) | 2 (0.9%) | 3 (0.7%) |  |
| 2 | 1 (0.9%) | 0 (0.0%) | 0 (0.0%) | 2 (0.9%) | 3 (0.7%) |  |
| 3 | 0 (0.0%) | 0 (0.0%) | 2 (3.4%) | 2 (0.9%) | 4 (0.9%) |  |
| 4 | 0 (0.0%) | 0 (0.0%) | 0 (0.0%) | 1 (0.4%) | 1 (0.2%) |  |
| 5 | 0 (0.0%) | 0 (0.0%) | 0 (0.0%) | 1 (0.4%) | 1 (0.2%) |  |
| Missing | 7 | 0 | 4 | 25 | 36 |  |
| **Hallucinations Severity**, n (%) |  |  |  |  |  | <.001^1^ |
| 0 | 109 (100.0%) | 24 (100.0%) | 53 (89.8%) | 226 (97.8%) | 412 (97.4%) |  |
| 1 | 0 (0.0%) | 0 (0.0%) | 5 (8.5%) | 5 (2.2%) | 10 (2.4%) |  |
| 2 | 0 (0.0%) | 0 (0.0%) | 1 (1.7%) | 0 (0.0%) | 1 (0.2%) |  |
| Missing | 7 | 0 | 4 | 25 | 36 |  |
| **Hallucinations Distress**, n (%) |  |  |  |  |  | <.001^1^ |
| 0 | 109 (100.0%) | 24 (100.0%) | 53 (89.8%) | 227 (98.3%) | 413 (97.6%) |  |
| 1 | 0 (0.0%) | 0 (0.0%) | 2 (3.4%) | 1 (0.4%) | 3 (0.7%) |  |
| 2 | 0 (0.0%) | 0 (0.0%) | 4 (6.8%) | 0 (0.0%) | 4 (0.9%) |  |
| 3 | 0 (0.0%) | 0 (0.0%) | 0 (0.0%) | 2 (0.9%) | 2 (0.5%) |  |
| 5 | 0 (0.0%) | 0 (0.0%) | 0 (0.0%) | 1 (0.4%) | 1 (0.2%) |  |
| Missing | 7 | 0 | 4 | 25 | 36 |  |
| **Agitation/Aggression Severity**, n (%) |  |  |  |  |  | .089^1^ |
| 0 | 97 (89.0%) | 22 (91.7%) | 49 (83.1%) | 183 (79.2%) | 351 (83.0%) |  |
| 1 | 8 (7.3%) | 0 (0.0%) | 7 (11.9%) | 24 (10.4%) | 39 (9.2%) |  |
| 2 | 4 (3.7%) | 2 (8.3%) | 3 (5.1%) | 24 (10.4%) | 33 (7.8%) |  |
| Missing | 7 | 0 | 4 | 25 | 36 |  |
| **Agitation/Aggression Distress**, n (%) |  |  |  |  |  | .079^1^ |
| 0 | 97 (89.0%) | 23 (95.8%) | 49 (83.1%) | 187 (81.0%) | 356 (84.2%) |  |
| 1 | 6 (5.5%) | 1 (4.2%) | 3 (5.1%) | 12 (5.2%) | 22 (5.2%) |  |
| 2 | 3 (2.8%) | 0 (0.0%) | 5 (8.5%) | 13 (5.6%) | 21 (5.0%) |  |
| 3 | 1 (0.9%) | 0 (0.0%) | 2 (3.4%) | 13 (5.6%) | 16 (3.8%) |  |
| 4 | 2 (1.8%) | 0 (0.0%) | 0 (0.0%) | 6 (2.6%) | 8 (1.9%) |  |
| Missing | 7 | 0 | 4 | 25 | 36 |  |
| **Dysphoria/Depression Severity**, n (%) |  |  |  |  |  | .007^1^ |
| 0 | 81 (74.3%) | 14 (58.3%) | 32 (54.2%) | 128 (55.4%) | 255 (60.3%) |  |
| 1 | 19 (17.4%) | 8 (33.3%) | 18 (30.5%) | 62 (26.8%) | 107 (25.3%) |  |
| 2 | 7 (6.4%) | 2 (8.3%) | 9 (15.3%) | 35 (15.2%) | 53 (12.5%) |  |
| 3 | 2 (1.8%) | 0 (0.0%) | 0 (0.0%) | 6 (2.6%) | 8 (1.9%) |  |
| Missing | 7 | 0 | 4 | 25 | 36 |  |
| **Dysphoria/Depression Distress**, n (%) |  |  |  |  |  | .024^1^ |
| 0 | 82 (75.2%) | 14 (58.3%) | 33 (55.9%) | 136 (59.1%) | 265 (62.8%) |  |
| 1 | 8 (7.3%) | 3 (12.5%) | 7 (11.9%) | 23 (10.0%) | 41 (9.7%) |  |
| 2 | 10 (9.2%) | 5 (20.8%) | 10 (16.9%) | 39 (17.0%) | 64 (15.2%) |  |
| 3 | 7 (6.4%) | 2 (8.3%) | 9 (15.3%) | 26 (11.3%) | 44 (10.4%) |  |
| 4 | 2 (1.8%) | 0 (0.0%) | 0 (0.0%) | 5 (2.2%) | 7 (1.7%) |  |
| 5 | 0 (0.0%) | 0 (0.0%) | 0 (0.0%) | 1 (0.4%) | 1 (0.2%) |  |
| Missing | 7 | 0 | 4 | 26 | 37 |  |
| **Anxiety Severity**, n (%) |  |  |  |  |  | .006^1^ |
| 0 | 86 (78.9%) | 17 (70.8%) | 43 (72.9%) | 143 (61.9%) | 289 (68.3%) |  |
| 1 | 17 (15.6%) | 6 (25.0%) | 10 (16.9%) | 48 (20.8%) | 81 (19.1%) |  |
| 2 | 5 (4.6%) | 1 (4.2%) | 6 (10.2%) | 32 (13.9%) | 44 (10.4%) |  |
| 3 | 1 (0.9%) | 0 (0.0%) | 0 (0.0%) | 8 (3.5%) | 9 (2.1%) |  |
| Missing | 7 | 0 | 4 | 25 | 36 |  |
| **Anxiety Distress**, n (%) |  |  |  |  |  | .002^1^ |
| 0 | 90 (82.6%) | 17 (70.8%) | 46 (78.0%) | 147 (63.6%) | 300 (70.9%) |  |
| 1 | 6 (5.5%) | 2 (8.3%) | 5 (8.5%) | 26 (11.3%) | 39 (9.2%) |  |
| 2 | 9 (8.3%) | 2 (8.3%) | 6 (10.2%) | 30 (13.0%) | 47 (11.1%) |  |
| 3 | 3 (2.8%) | 3 (12.5%) | 2 (3.4%) | 18 (7.8%) | 26 (6.1%) |  |
| 4 | 1 (0.9%) | 0 (0.0%) | 0 (0.0%) | 7 (3.0%) | 8 (1.9%) |  |
| 5 | 0 (0.0%) | 0 (0.0%) | 0 (0.0%) | 3 (1.3%) | 3 (0.7%) |  |
| Missing | 7 | 0 | 4 | 25 | 36 |  |
| **Euphoria/Elation Severity**, n (%) |  |  |  |  |  | .721^1^ |
| 0 | 103 (95.4%) | 24 (100.0%) | 57 (96.6%) | 220 (95.2%) | 404 (95.7%) |  |
| 1 | 3 (2.8%) | 0 (0.0%) | 0 (0.0%) | 5 (2.2%) | 8 (1.9%) |  |
| 2 | 2 (1.9%) | 0 (0.0%) | 2 (3.4%) | 4 (1.7%) | 8 (1.9%) |  |
| 3 | 0 (0.0%) | 0 (0.0%) | 0 (0.0%) | 2 (0.9%) | 2 (0.5%) |  |
| Missing | 8 | 0 | 4 | 25 | 37 |  |
| **Euphoria/Elation Distress**, n (%) |  |  |  |  |  | .817^1^ |
| 0 | 105 (97.2%) | 24 (100.0%) | 57 (96.6%) | 225 (97.8%) | 411 (97.6%) |  |
| 1 | 3 (2.8%) | 0 (0.0%) | 2 (3.4%) | 2 (0.9%) | 7 (1.7%) |  |
| 2 | 0 (0.0%) | 0 (0.0%) | 0 (0.0%) | 1 (0.4%) | 1 (0.2%) |  |
| 3 | 0 (0.0%) | 0 (0.0%) | 0 (0.0%) | 1 (0.4%) | 1 (0.2%) |  |
| 4 | 0 (0.0%) | 0 (0.0%) | 0 (0.0%) | 1 (0.4%) | 1 (0.2%) |  |
| Missing | 8 | 0 | 4 | 26 | 38 |  |
| **Apathy/Indifference Severity**, n (%) |  |  |  |  |  | <.001^1^ |
| 0 | 97 (89.0%) | 23 (95.8%) | 44 (74.6%) | 170 (73.6%) | 334 (79.0%) |  |
| 1 | 9 (8.3%) | 1 (4.2%) | 7 (11.9%) | 27 (11.7%) | 44 (10.4%) |  |
| 2 | 2 (1.8%) | 0 (0.0%) | 8 (13.6%) | 27 (11.7%) | 37 (8.7%) |  |
| 3 | 1 (0.9%) | 0 (0.0%) | 0 (0.0%) | 7 (3.0%) | 8 (1.9%) |  |
| Missing | 7 | 0 | 4 | 25 | 36 |  |
| **Apathy/Indifference Distress**, n (%) |  |  |  |  |  | .007^1^ |
| 0 | 98 (89.9%) | 23 (95.8%) | 45 (76.3%) | 178 (77.1%) | 344 (81.3%) |  |
| 1 | 4 (3.7%) | 0 (0.0%) | 5 (8.5%) | 16 (6.9%) | 25 (5.9%) |  |
| 2 | 3 (2.8%) | 1 (4.2%) | 5 (8.5%) | 20 (8.7%) | 29 (6.9%) |  |
| 3 | 3 (2.8%) | 0 (0.0%) | 4 (6.8%) | 13 (5.6%) | 20 (4.7%) |  |
| 4 | 1 (0.9%) | 0 (0.0%) | 0 (0.0%) | 2 (0.9%) | 3 (0.7%) |  |
| 5 | 0 (0.0%) | 0 (0.0%) | 0 (0.0%) | 2 (0.9%) | 2 (0.5%) |  |
| Missing | 7 | 0 | 4 | 25 | 36 |  |
| **Disinhibition Severity**, n (%) |  |  |  |  |  | .047^1^ |
| 0 | 99 (90.8%) | 23 (95.8%) | 48 (81.4%) | 187 (81.0%) | 357 (84.4%) |  |
| 1 | 3 (2.8%) | 1 (4.2%) | 7 (11.9%) | 21 (9.1%) | 32 (7.6%) |  |
| 2 | 5 (4.6%) | 0 (0.0%) | 3 (5.1%) | 19 (8.2%) | 27 (6.4%) |  |
| 3 | 2 (1.8%) | 0 (0.0%) | 1 (1.7%) | 4 (1.7%) | 7 (1.7%) |  |
| Missing | 7 | 0 | 4 | 25 | 36 |  |
| **Disinhibition Distress**, n (%) |  |  |  |  |  | .431^1^ |
| 0 | 100 (91.7%) | 23 (95.8%) | 52 (88.1%) | 201 (87.0%) | 376 (88.9%) |  |
| 1 | 2 (1.8%) | 0 (0.0%) | 1 (1.7%) | 10 (4.3%) | 13 (3.1%) |  |
| 2 | 3 (2.8%) | 1 (4.2%) | 2 (3.4%) | 11 (4.8%) | 17 (4.0%) |  |
| 3 | 1 (0.9%) | 0 (0.0%) | 4 (6.8%) | 6 (2.6%) | 11 (2.6%) |  |
| 4 | 3 (2.8%) | 0 (0.0%) | 0 (0.0%) | 3 (1.3%) | 6 (1.4%) |  |
| Missing | 7 | 0 | 4 | 25 | 36 |  |
| **Irritability/Lability Severity**, n (%) |  |  |  |  |  | <.001^1^ |
| 0 | 88 (80.7%) | 18 (75.0%) | 36 (61.0%) | 136 (58.9%) | 278 (65.7%) |  |
| 1 | 17 (15.6%) | 3 (12.5%) | 14 (23.7%) | 48 (20.8%) | 82 (19.4%) |  |
| 2 | 3 (2.8%) | 2 (8.3%) | 9 (15.3%) | 33 (14.3%) | 47 (11.1%) |  |
| 3 | 1 (0.9%) | 1 (4.2%) | 0 (0.0%) | 14 (6.1%) | 16 (3.8%) |  |
| Missing | 7 | 0 | 4 | 25 | 36 |  |
| **Irritability/Lability Distress**, n (%) |  |  |  |  |  | .014^1^ |
| 0 | 89 (81.7%) | 19 (79.2%) | 40 (67.8%) | 150 (65.2%) | 298 (70.6%) |  |
| 1 | 7 (6.4%) | 1 (4.2%) | 7 (11.9%) | 28 (12.2%) | 43 (10.2%) |  |
| 2 | 7 (6.4%) | 2 (8.3%) | 6 (10.2%) | 18 (7.8%) | 33 (7.8%) |  |
| 3 | 4 (3.7%) | 1 (4.2%) | 6 (10.2%) | 25 (10.9%) | 36 (8.5%) |  |
| 4 | 2 (1.8%) | 1 (4.2%) | 0 (0.0%) | 7 (3.0%) | 10 (2.4%) |  |
| 5 | 0 (0.0%) | 0 (0.0%) | 0 (0.0%) | 2 (0.9%) | 2 (0.5%) |  |
| Missing | 7 | 0 | 4 | 26 | 37 |  |
| **Aberrant Motor Severity**, n (%) |  |  |  |  |  | .272^1^ |
| 0 | 102 (93.6%) | 22 (91.7%) | 54 (91.5%) | 201 (87.0%) | 379 (89.6%) |  |
| 1 | 5 (4.6%) | 2 (8.3%) | 2 (3.4%) | 19 (8.2%) | 28 (6.6%) |  |
| 2 | 1 (0.9%) | 0 (0.0%) | 3 (5.1%) | 6 (2.6%) | 10 (2.4%) |  |
| 3 | 1 (0.9%) | 0 (0.0%) | 0 (0.0%) | 5 (2.2%) | 6 (1.4%) |  |
| Missing | 7 | 0 | 4 | 25 | 36 |  |
| **Aberrant Motor Distress**, n (%) |  |  |  |  |  | .286^1^ |
| 0 | 106 (97.2%) | 23 (95.8%) | 54 (91.5%) | 213 (92.2%) | 396 (93.6%) |  |
| 1 | 1 (0.9%) | 1 (4.2%) | 2 (3.4%) | 6 (2.6%) | 10 (2.4%) |  |
| 2 | 0 (0.0%) | 0 (0.0%) | 2 (3.4%) | 5 (2.2%) | 7 (1.7%) |  |
| 3 | 2 (1.8%) | 0 (0.0%) | 0 (0.0%) | 6 (2.6%) | 8 (1.9%) |  |
| 4 | 0 (0.0%) | 0 (0.0%) | 1 (1.7%) | 1 (0.4%) | 2 (0.5%) |  |
| Missing | 7 | 0 | 4 | 25 | 36 |  |
| **Nighttime Behavior Severity**, n (%) |  |  |  |  |  | .004^1^ |
| 0 | 91 (83.5%) | 14 (58.3%) | 42 (71.2%) | 150 (64.9%) | 297 (70.2%) |  |
| 1 | 6 (5.5%) | 5 (20.8%) | 5 (8.5%) | 25 (10.8%) | 41 (9.7%) |  |
| 2 | 10 (9.2%) | 4 (16.7%) | 10 (16.9%) | 41 (17.7%) | 65 (15.4%) |  |
| 3 | 2 (1.8%) | 1 (4.2%) | 2 (3.4%) | 15 (6.5%) | 20 (4.7%) |  |
| Missing | 7 | 0 | 4 | 25 | 36 |  |
| **Nighttime Behavior Distress**, n (%) |  |  |  |  |  | .182^1^ |
| 0 | 92 (84.4%) | 20 (83.3%) | 44 (74.6%) | 173 (74.9%) | 329 (77.8%) |  |
| 1 | 4 (3.7%) | 1 (4.2%) | 5 (8.5%) | 14 (6.1%) | 24 (5.7%) |  |
| 2 | 8 (7.3%) | 1 (4.2%) | 6 (10.2%) | 17 (7.4%) | 32 (7.6%) |  |
| 3 | 4 (3.7%) | 2 (8.3%) | 4 (6.8%) | 19 (8.2%) | 29 (6.9%) |  |
| 4 | 0 (0.0%) | 0 (0.0%) | 0 (0.0%) | 6 (2.6%) | 6 (1.4%) |  |
| 5 | 1 (0.9%) | 0 (0.0%) | 0 (0.0%) | 2 (0.9%) | 3 (0.7%) |  |
| Missing | 7 | 0 | 4 | 25 | 36 |  |
| **Appetite/Eating Severity**, n (%) |  |  |  |  |  | .017^1^ |
| 0 | 92 (84.4%) | 19 (79.2%) | 39 (66.1%) | 163 (70.6%) | 313 (74.0%) |  |
| 1 | 12 (11.0%) | 3 (12.5%) | 13 (22.0%) | 37 (16.0%) | 65 (15.4%) |  |
| 2 | 4 (3.7%) | 2 (8.3%) | 5 (8.5%) | 24 (10.4%) | 35 (8.3%) |  |
| 3 | 1 (0.9%) | 0 (0.0%) | 2 (3.4%) | 7 (3.0%) | 10 (2.4%) |  |
| Missing | 7 | 0 | 4 | 25 | 36 |  |
| **Appetite/Eating Distress**, n (%) |  |  |  |  |  | .014^1^ |
| 0 | 100 (91.7%) | 22 (91.7%) | 48 (81.4%) | 181 (78.4%) | 351 (83.0%) |  |
| 1 | 2 (1.8%) | 1 (4.2%) | 2 (3.4%) | 15 (6.5%) | 20 (4.7%) |  |
| 2 | 3 (2.8%) | 1 (4.2%) | 4 (6.8%) | 16 (6.9%) | 24 (5.7%) |  |
| 3 | 3 (2.8%) | 0 (0.0%) | 5 (8.5%) | 13 (5.6%) | 21 (5.0%) |  |
| 4 | 1 (0.9%) | 0 (0.0%) | 0 (0.0%) | 5 (2.2%) | 6 (1.4%) |  |
| 5 | 0 (0.0%) | 0 (0.0%) | 0 (0.0%) | 1 (0.4%) | 1 (0.2%) |  |
| Missing | 7 | 0 | 4 | 25 | 36 |  |
| **NPI-Q Severity Score** |  |  |  |  |  | <.001^1^ |
| N (Missing) | 108 (8) | 24 (0) | 59 (4) | 231 (25) | 422 (37) |  |
| Mean (SD) | 2.0 (3.3) | 2.5 (2.3) | 3.8 (4.5) | 4.4 (4.8) | 3.6 (4.4) |  |
| Median (IQR) | 1.0 (0.0, 3.0) | 2.0 (0.5, 4.0) | 2.0 (0.0, 6.0) | 3.0 (0.0, 7.0) | 2.0 (0.0, 6.0) |  |
| Median (Range) | 1.0 (0.0, 14.0) | 2.0 (0.0, 8.0) | 2.0 (0.0, 20.0) | 3.0 (0.0, 23.0) | 2.0 (0.0, 23.0) |  |
| **NPI-Q Distress Score** |  |  |  |  |  | <.001^1^ |
| N (Missing) | 108 (8) | 24 (0) | 59 (4) | 230 (26) | 421 (38) |  |
| Mean (SD) | 2.5 (4.7) | 2.7 (3.9) | 4.4 (5.9) | 5.1 (6.3) | 4.2 (5.9) |  |
| Median (IQR) | 0.0 (0.0, 3.0) | 1.0 (0.0, 4.0) | 1.0 (0.0, 8.0) | 2.0 (0.0, 8.0) | 2.0 (0.0, 6.0) |  |
| Median (Range) | 0.0 (0.0, 22.0) | 1.0 (0.0, 15.0) | 1.0 (0.0, 23.0) | 2.0 (0.0, 32.0) | 2.0 (0.0, 32.0) |  |
| ^1^Kruskal-Wallis *p*-value; | | | | | | |
