## Supplementary figures and images for "Neuropsychiatric Symptom Associations with *NOTCH3* Genotype in the United States CADASIL Consortium"

### Supplemental Figure 1

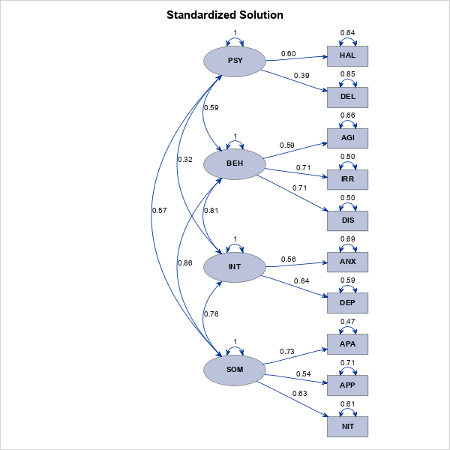
